# MaTiLDA: An Integrated Machine Learning and Topological Data Analysis Platform for Brain Network Dynamics

**DOI:** 10.1101/2023.06.08.23290830

**Authors:** Katrina Prantzalos, Dipak Upadhyaya, Nassim Shafiabadi, Nick Gurski, Guadalupe Fernandez-BacaVaca, Kenneth Yoshimoto, Subhashini Sivagnanam, Amitava Majumdar, Satya S. Sahoo

**Affiliations:** Department of Population and Quantitative Health Sciences, Case Western Reserve University School of Medicine, Cleveland, OH, USA; Department of Mathematics, Case Western Reserve University, Cleveland, OH, USA; Department of Neurology, University Hospitals Cleveland Medical Center, Cleveland, OH, USA; San Diego Supercomputer Center, University of California, San Diego, CA, USA

## Abstract

Topological data analysis (TDA) is a powerful approach for investigating complex relationships in brain networks; however, its application requires substantial domain knowledge in programming, mathematics, and data science, especially in the context of data-driven approaches like machine learning (ML). To address this educational barrier, we introduce MaTiLDA, a graphical user interface that enables exploration of common representations of TDA features and their efficacy in various classical machine learning models. This user-friendly tool is the first graphical user interface built to explore TDA representations in machine learning applications. MaTiLDA provides a user-centric method for characterizing complex neural relationships using TDA techniques. To demonstrate the utility of MaTiLDA in characterizing brain network dynamics, we apply this workflow to a cohort of 4 refractory epilepsy patients and evaluate the predictive performance of various TDA feature representations in a series of ML models.

The MaTiLDA application can be accessed through https://bmhinformatics.case.edu/nic/MaTiLDA

## 1. Introduction

The increasing availability of multimodal brain network data emphasizes an emergent demand for accurate and reliable analytic methods to characterize these networks to meet critical clinical objectives and improve patient care^1^. Networks within the brain, such as the default mode network, the salience network, and the central-executive network, provide the physiological basis for a wide range of cognitive functions^2^. Understanding disruptions in these networks is crucial to characterizing neurological disorders, revealing pathophysiological mechanisms, and defining biomarkers for clinical diagnoses^1,3,4^. For example, characterizing the dynamics of epilepsy seizure networks is an important clinical objective of translational research in epilepsy. Epilepsy is the second most common neurological disorder, affecting over 50 million individuals worldwide^5^. Epilepsy is characterized by recurrent seizures stemming from abnormal electrical discharges which spread throughout the brain^5,6^. Similar to other disease domains, there has been a rapid increase in the use of machine learning (ML) algorithms in studying brain network dynamics in epilepsy patients^7–9^. ML algorithms such as Support Vector Machine (SVM) methods have used features extracted from signal processing methods applied to electroencephalogram (EEG) recording to accurately lateralize brain regions responsible for seizure onset^10–12^. Recent studies have derived algebraic topology structures from EEG data to address several limitations of graph model based analysis of brain networks^13–15^. Topological data analysis, a mathematical framework that leverages tools from algebraic topology, enables a quantitative analysis of higher-dimensional interactions in complex data by using robust scale-invariant methods to infer relevant information regarding the topological structure of the data^16,17^.

In the past decade, topological data analysis has seen progress in brain network analysis using techniques such as persistent homology, particularly in the domain of brain network analysis^13,15,18–21^. Specifically, statistical summaries of results from persistent homology, such as persistence landscapes, persistence images, and persistent entropy have highlighted the promise of topological data analysis in identifying regions of seizure activity^19,21^ and distinguishing seizure onset from preictal activity^13,18^. Findings demonstrate similarities between persistence landscapes representing brain activity at the seizure origin site during seizure and pre-seizure periods; however, persistence landscapes from pre-seizure periods are evidenced to exhibit stability whereas persistence landscapes from seizure periods are characterized by the dominance of a single topological structure^19,21^. Alternatively, a study of persistent entropy demonstrated that seizure activity is characterized by topological structures with increased persistence, whereas activity from pre-seizure periods demonstrates a higher prevalence of topological structures and a wider range in their persistence^18^. Although there is variation in defining the characteristics of topological structures to distinguish seizure and pre-seizure activity, it is evident that TDA measures consistently excel in distinguishing aberrant brain activity^13,18–21^. However, it is essential to acknowledge that the development and utilization of a machine learning and topological data analysis framework to achieve these results is a resource-intensive endeavor that demands substantial expertise in fields such as machine learning, computer science, and data analysis. While many programming tools exist to simplify topological data analysis and machine learning, such as GUDHI, RIPSER, DIONYSUS, PERSEUS, JHOLES, and JAVAPLEX, these tools still require a large amount of knowledge in programming and data analysis before any actionable insights can be derived^22^. Obtaining expertise in these skillsets can be challenging given the substantial time and effort required, creating a significant barrier to entry for smaller labs, especially those in non-computational fields. Thus, the application of ML using TDA features is extremely challenging for the wider neuroscience research community who have limited experience in both TDA and ML algorithm implementation.

To address this critical barrier to the integrated adoption of ML together with TDA to analyze the increasingly large volumes of brain network EEG recording, we introduce MaTiLDA, an interface to enable systematic ML analysis of topological structures derived from signal data. In MaTiLDA, we deploy a user-friendly web application to host our framework for classifying dynamic brain states using machine learning analyses of features from topological data analysis applied to EEG. We aim to facilitate the investigation of topological structures for characterizing anomalous brain network activity.

### 1.1 Background

#### The Neuro-Integrative Connectivity Workflow

Over the past decade, we have developed an integrated informatics platform, named the Neuro-Integrative Connectivity (NIC) tool, designed for the systematic characterization of brain network dynamics in epilepsy patients^14,23,24^. The fundamental objective of the NIC tool is to alleviate the burdensome tasks faced by researchers and clinicians when processing EEG data. NIC aims to reduce the substantial challenges associated with data management in signal processing by offering a workflow that simplifies the complexities of multi-step methodologies for preprocessing and analyzing signal data across numerous seizure events. Currently, the NIC tool is comprised of four modules: the converter, the correlator, the network analysis module, and the topological data analysis module. These modules are used to convert signal data from European Data Format (EDF) into a format more suitable for storage, sharing, and analysis, to compute signal coupling in EEG data, and to analyze this signal coupling with common network analysis and topological data analysis measures. The converter processes signal data stored in EDF files into Cloudwave Signal Format (CSF) files, a JavaScript Object Notation (JSON) based human-readable format with semantic annotations using epilepsy domain ontology^23^. The correlator leverages common correlation metrics, Pearson linear correlation^25^, mean phase coherence^26^, and the nonlinear regression coefficient *h*^*2*^ by Pijn et al.^27^, to characterize the association between two EEG signals. The network analysis and topological data analysis modules enable the calculation of common network analysis metrics and the use of persistent homology, respectively. We refer to our previous work^14,23,24^ for more information on the NIC tool, which can be download at https://bmhinformatics.case.edu/nicworkflow. The application proposed in this study, MaTiLDA, is an extension of the NIC workflow that will allow users to conduct machine learning analysis of topological structures computed from the signal coupling data that the NIC tool collects from EEG data.

#### Topological Features from Stereotactic Electroencephalography Signal Coupling

Stereotactic EEG (SEEG) is a high-resolution imaging modality that records signal data from multi-contact electrodes implanted directly in brain structures^6^. In epilepsy, the changes in SEEG signals provide insight into how local network variations affect global changes in brain activity^28^, and these signals are widely regarded as the gold standard in pre-surgical evaluation for identifying the critical area for surgical resection in epilepsy patients^29^. SEEG records interactions between electrodes; these interactions can be characterized via signal coupling in various brain regions^14^. The NIC tool allows users to derive this signal coupling from raw EEG using various correlation metrics, storing this information in a correlation matrix^14^. Persistent homology, a topological data analysis method that tracks the formation (birth) and termination (death) of topological features across a filtration, can be applied to matrices of signal coupling to identify multidimensional interactions and reveal changes in topological structures underpinning the breakdown of normal dynamic brain networks^22,28,30^. In this process, a filtration is applied to a correlation matrix representing the signal coupling in SEEG data, gradually varying the scale or threshold to extract topological features at different levels of connectivity^13,22^. The persistence of these features, measured by their creation (birth) and termination (death), is then analyzed to identify changes in topological structures and gain insights into the topology of brain networks^17,22,30,31^. Theoretical foundations of these methods are described by Hatcher^17^ and Edelsbrunner et al.^30^.

#### Topological Feature Representation in Machine Learning

The results from persistent homology analysis pose challenges for direct utilization in statistical or machine learning analysis^32^. Persistent homology tracks the birth and death of topological features and the resulting multiset of birth, death tuples is commonly visualized with a persistence diagram, a plot representing birth filtration along the x axis and death filtration along the y axis^22,30,33^. Persistence diagrams are powerful tools for quantifying shape in geometric data, as they are resilient to noise-induced perturbations, offering robustness and stability^34^. However, several aspects of persistence diagrams make them difficult to use directly in machine learning analyses and statistical comparisons. The inconsistency in the number of points across diagrams creates a challenge for conducting balanced comparisons. Moreover, persistence diagrams are not vectors in a Hilbert or Banach space and thus a unique mean cannot be established to define statistical measures^22,35^. Undoubtedly, the direct utilization of persistence diagrams in machine learning models poses significant challenges^32^. Consequently, a range of vectorization methods have been devised to facilitate the integration of persistent homology features into machine learning classifications. Vectorization, also known as featurization, is the process of mapping a persistence diagram to Euclidean space^33^. Numerous vectorization methods have been proposed —including persistence landscapes, persistence images, and persistent entropy— and this area of research is rapidly advancing^33^. In this work, we show how MaTiLDA can be used to intuitively apply various persistence diagram vectorization methods with a sample of classical machine learning algorithms.

## 2. Methods

The computation and comparative analysis of topological features from raw signal data related to clinical events entails multiple stages of data manipulation and analysis, which include extraction of signal data, computation of signal coupling, topological data analysis of signal coupling, data cleaning, and comparative analysis of topological features (Figure 1). Scientific workflow systems, like the NIC workflow previously established by our lab, have been used to automate these multi-step process^14^. In this work, we describe a continuation of the NIC workflow that incorporates tools from topological data analysis into a modular application for brain network analysis. In this section, we introduce MaTiLDA, a machine learning pipeline for processing features from various vectorization methods of results from persistent homology analysis of SEEG data. The primary goals of MaTiLDA are:

1. To provide an accessible tool for analyzing neural relationships
2. To develop a platform that enables topological data analysis without requiring extensive technical expertise
3. To provide an efficient approach for optimizing brain network analysis using machine learning and topological data analysis

**Figure 1:**
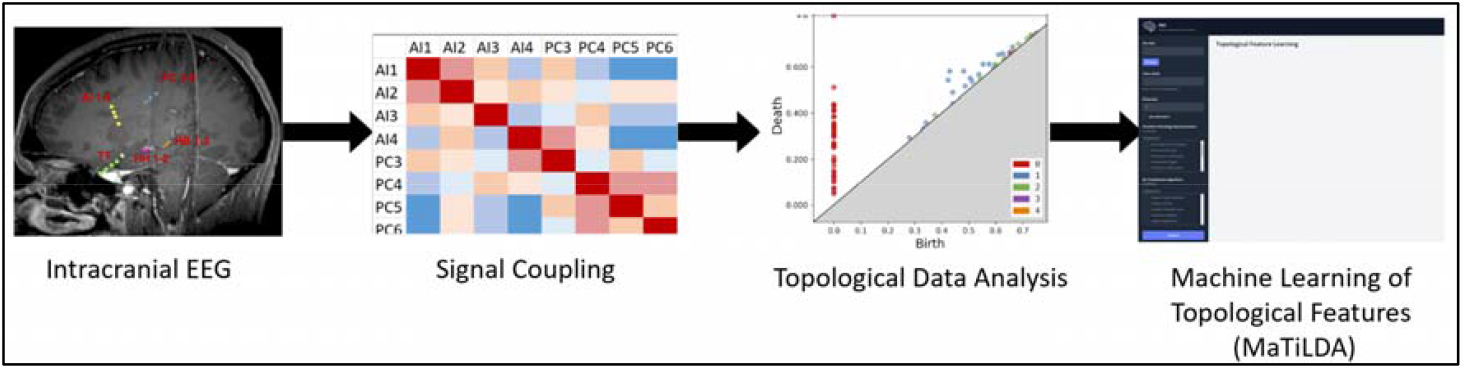
Our framework for computing and comparing topological features from dynamic brain networks. Stereotactic EEG is used to extract signal data during epileptic seizures. Signal coupling between various brain regions active during a seizure is calculated using a nonlinear correlation coefficient. The signal coupling matrices are processed using a Vietoris-Rips filtration. The (birth, death) pairs describing the resulting topological features are then vectorized and used as input into an analysis method such as a Support Vector Machine.

### 2.1 A Framework for Classifying Brain States

The MaTiLDA tool developed in this work is a visual interface for working with a version of a framework previously developed by our lab (Figure 2)^14^. We leverage the converter and correlator from the NIC workflow to convert EDF files from SEEG signal data and generate matrices of signal coupling. MaTiLDA is passed a series of signal coupling matrices and applies a Vietoris-Rips filtration to each matrix using methods from the GUDHI^36^ library for topological data analysis to generate persistence diagrams. The MaTiLDA user interface allows users to select a series of vectorizations from GUDHI’s library for topological data analysis. The vectorizations are then used, based on user model selection, in classical machine learning models from various Scikit-learn functions.

**Figure 2:**
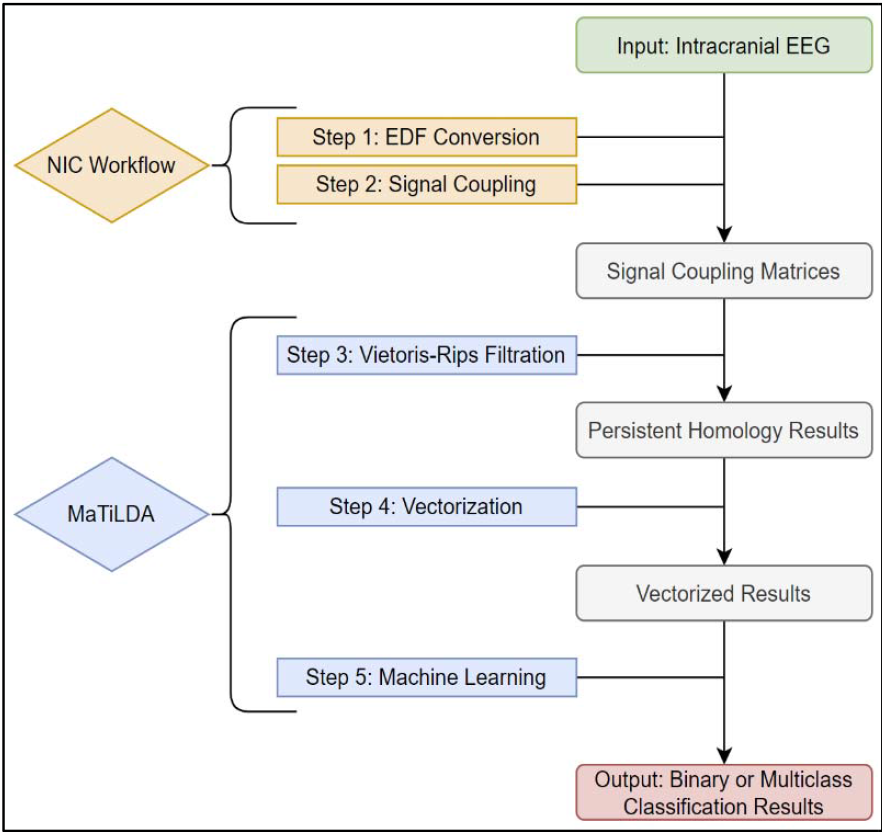
The MaTiLDA workflow leverages the NIC workflow to create matrices of signal coupling. We apply a Rips filtration to these matrices to get persistence diagrams that are vectorized for machine learning analysis based on user inputs.

### 2.2 MaTiLDA Architecture and Development

For MaTiLDA, a simple graphical user interface was built using a Django web application framework, which uses the Python programming language and features several libraries and modules that support a variety of data processing and analysis tasks including libraries for machine learning and topological data analysis. The MaTiLDA platform uses the Model View Template (MVT) approach with data inputs being managed using an object relational data Model, the user interface is managed by the View component, and the user interaction with various features of the software is mediated by the Template, which conforms to the Django framework. The MaTiLDA platform is accessed via a web browser with role-based access control (RBAC) with users assigned to a user group. Users follow an intuitive process to conduct various analyses of their data. Results are generated upon clicking the “Analyze Data” button.

The steps of the MaTiLDA pipeline are outlined in ***Figure 2***. Using the NIC tool, users may process iEEG data to create matrices of signal coupling. We use these matrices of signal coupling as input to MaTiLDA. MaTiLDA requests a folder containing a set of these coupling matrices, the labels with which to classify them, a dimension for analysis, a list of vectorization methods to apply, and a list of machine learning algorithms to run. MaTiLDA takes each of the provided files and applies a Vietoris-Rips filtration using GUDHI’s persistent homology toolkit. For each file, persistent homology results are vectorized as requested by the user. The vector is assigned a label based on the associated file path matched to the provided list of class labels. It is assumed that at all signal coupling matrix files belonging to the desired class are contained in a folder named the same thing as the desired class label; this folder should be a subfolder of the provided file path. All vectorization methods are implemented using methods from the GUDHI library for topological data analysis with default parameters. An 80/20 train-test split is applied to the vectorized dataset, and a machine learning model is trained using the 80% data partition. Labels are predicted for the 20% test set, and the test set accuracy score is reported alongside the precision, recall, and the area under the receiver operating curve (AUC). For multiclass models, precision, recall, and AUC are calculated using the one-versus-all approach implemented in Scikit-learn. Note that a separate machine learning model is run for each algorithm for each vectorization. For example, if a user selects 3 vectorization methods and 4 machine learning algorithms, a total of 12 models will be run. All machine learning models are implemented using the Scikit-learn library with default model parameters.

### 2.3 MaTiLDA User Interface

The MaTiLDA user interface (***Figure 3***) consists of an intuitive data entry module and a minimal results table (***Figure 4***). The architecture of MaTiLDA allows users to specify a file location containing files previously processed using the NIC’s Correlator tool. It is expected that this file location will contain subfolders with names matching class labels that will be provided by the user; each subfolder should contain the data for that specific label. The user must specify a dimension for which they would like to analyze data; however, the user may opt to set the given dimension as the max dimension for analysis and consider all dimensions up to and including this dimension in their analysis. Users may select several vectorization methods and machine learning algorithms from a set of checkbox lists. Upon clicking the “Analyze” button, results will be generated for all vector-model pair selected. The results table along the right side of the screen shows the vectorization chosen, the machine learning algorithm used, the model’s accuracy in testing data, the true positive rate, the false negative rate, and the area under the receiver operating curve.

**Figure 3:**
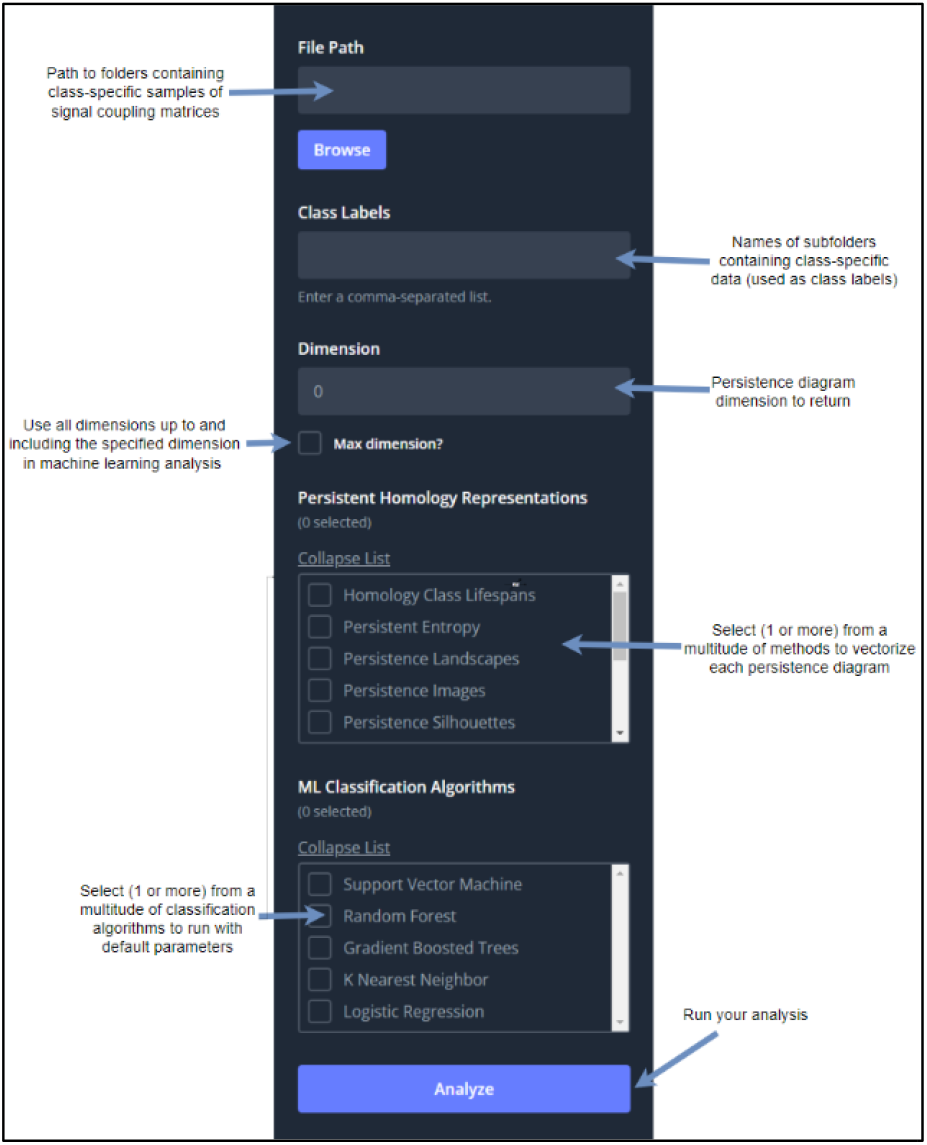
MaTiLDA supports various vectorizations and ML methods.

**Figure 4:**
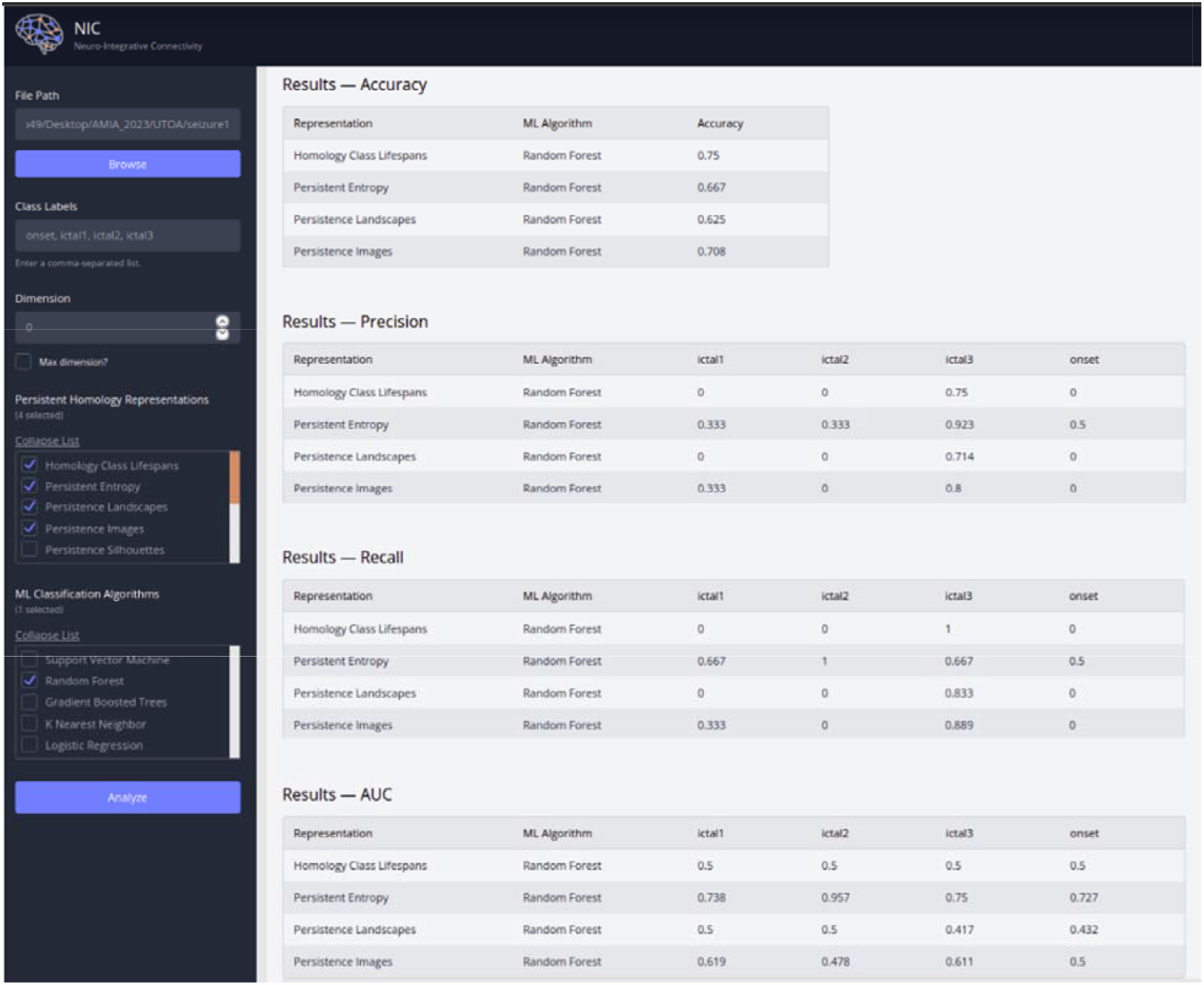
Results for one seizure from a multiclass classification of seizure periods using Homology Class Lifespan vectors, persistent entropy vectors, persistence landscape vectors, or persistence image vectors as input to a Random Forest model.

### 2.4 Topological Data Analysis of SEEG

The MaTiLDA application is a continuation of the modular NIC workflow. In the MaTiLDA pipeline (***Figure 2***), raw EEG data is converted to signal coupling matrices, which are processed using persistent homology. The persistent homology results are vectorized based on user specifications and the resulting vectors are used as input machine learning models. In this section, we describe the vectorization methods available to users through MaTiLDA.

#### Homology Class Lifespans

We calculate the lifespan, death minus birth, for each (birth, death) pair describing the persistence of a topological structure as a result of persistent homology. These lifespan values are stored in a vector such that there is one vector per set of results from persistent homology. For any vectors not matching the length of the longest vector in the full dataset, we pad with zero. These methods mimic those outlined by Bendich et al.^37^; however, unlike Bendich et al., we do not limit the number of lifespan values included in a vector.

#### Persistence Landscapes & Silhouettes

Persistence landscapes transform persistence diagrams into a Hilbert space by rotating a persistence diagram such that the y=x diagonal becomes the new x-axis and applying a tent function, *f*_*a,b*_(*t*) = max(0, min (*a* + *t, b* − *t*)), to each point on the diagram *D* = (*a*_*i*_, *b*_*i*_)_*i∈I*_.^38^ A vector is created by uniformly sampling values along the new x-axis and calculating *λ*_*k*_ (*t*) = *kmax*{ *f*_*a,b*_(*t*)}_*i∈I*_ at each x-value^38,39^. A persistence silhouette is a variation of the persistence landscape in which a vector is created by taking the weighted average of the tent functions, rather than the maximum^40^.

#### Persistence Images

Persistence images convert a set of birth-lifespan pairs resulting from persistent homology into a two-dimensional image where each pixel represents a rectangular area of the diagram, and the intensity of the image represents the frequency of occurrence of the birth-lifespan pair. These images can then be flattened into a vector with the various pixel values of the persistence image. The resolution of the image, thus, determines the number of features computed by the persistent image^41^.

#### Persistent Entropy

Persistent entropy is described as the Shannon entropy of the persistence of topological structures. The persistent entropy is calculated using the function 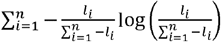 where l_i_ is the lifespan of a topological structure^42^.

### 2.5 Classical Machine Learning of TDA Features

Six common classical algorithms for machine learning classification were selected to be implemented in the initial version of MaTiLDA: support vector machines, random forest, gradient boosted trees, K-nearest neighbor, and logistic regression. We provide a brief introduction to each algorithm here.

#### Support Vector Machine

Support Vector Machine (SVM) is a supervised learning algorithm that aims to find the best-separating hyperplane to maximize the margin between two classes to classify data into different categories ^43^.To do this, SVM uses a kernel function to map the input data to a higher-dimensional feature space where it can be more easily separated. SVM then finds the optimal hyperplane that separates the classes by maximizing the distance between the hyperplane and the closest points from each class, called support vectors^44^. In the case of non-linearly separable data, SVM uses a kernel to transform the data into a higher-dimensional space that is linearly separable. SVM has several advantages over classical classification algorithms, such as logistic regression and decision trees, as it is less prone to overfitting, works well with high-dimensional data, and can handle both binary and multi-class classification problems^43,44^.

#### Random Forest & Gradient Boosted Trees

Random forest is a widely used ensemble learning algorithm that combines multiple decision trees to make predictions. It has gained popularity in machine learning due to its high classification and regression task accuracy^45^. The algorithm’s fundamental concept is to mitigate overfitting and enhance the model’s accuracy by introducing randomness, which is accomplished by training each tree on a random subset of data. Each tree is created by recursively dividing the data into smaller subsets based on feature values to maximize information gain or minimize subset impurity. A majority vote from the tree-specific predictions is used to define the final classification label; the final prediction is the category that receives the most votes^45,46^. Gradient boosted trees (GBT) are a similar method to random forest. GBT is a powerful sequential learning algorithm that can learn complex, non-linear relationships between features and the target variable. Where RF constructs an ensemble of decision trees independently, each tree in GBT is constructed to correct mistakes of the previous trees based on previous misclassifications. Its main advantage over RF is its ability to model complex non-linear relationships, while its main disadvantage is its potential for overfitting^45,46^.

#### K-Nearest Neighbor

K-nearest neighbor (KNN) is a non-parametric, supervised learning classifier that facilitates binary or multiclass classification for unlabeled observations by leveraging their proximity to the K nearest datapoints, or neighbors, in the training data. The classification decision is made through a majority voting scheme among the K nearest neighbors. While KNN offers an intuitive interpretability, it is accompanied by a significant computational cost due to the necessity of performing distance calculations for each new observation.^43,45,46^.

#### Logistic Regression

Logistic regression (LR) is a statistical learning method used for binary classification. By employing a logistic function, 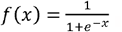, a linear combination of predictors is mapped to the range [0, 1], allowing LR to estimate the probability of the binary outcome using maximum likelihood estimation. LR is a low-complexity model with highly interpretable results; however, it assumes a linear relationship between predictors and the log-odds of the outcome, is sensitive to outliers, and can be prone to overfitting^43,44,46^.

### 2.6 Study Data

We randomly selected two seizures each from four refractory epilepsy patients receiving care in the epilepsy monitoring unit (EMU) at University Hospitals Cleveland Medical Center’s Tier 4 epilepsy center. ***Table 1*** shows the characteristics of these patients and their seizures. EMU patients are continuously monitored with SEEG for 5-10 days to evaluate surgical resection of the epileptogenic zone. Intracranial electrodes are placed based on a presurgical protocol described elsewhere^47,48^. Retrospective visual analyses of SEEG recordings were conducted using a Nihon-Kohden Neurofax system (Nihon Kohden America, Foothill Ranch, CA, U.S.A.) with AC amplifiers, a high sampling rate of 2,000 Hz, and an acquisition rate spanning 0.016-300 Hz. The SEEG was filtered at 600 Hz with a 0.03s time constant and sensitivity ranging from 30-100 μV based on optimal seizure visibility for each implant. A 60 Hz notch filter was applied universally. Clinicians defined seizure onset as the earliest distinctive occurrence of rhythmic sinusoidal activity or repetitive spikes; the region of activity was noted as the seizure onset zone^49^. Seizure phases were defined as the subsequent spread of seizure activity. EEG sequences were broken down into one second epochs and features were computed for each epoch.

**Table 1:** Characteristics of two seizures from four randomly selected refractory epilepsy patients.

| Patient | Age Range | Sex | Epileptogenic Zone | Medication | Seizure Duration (s) | Active Electrodes | Seizure Phases | Seizure Semiology |
| --- | --- | --- | --- | --- | --- | --- | --- | --- |
| 1 | 25-30 | F | Left Hemisphere | Trileptal, Keppra | 48 | IM1, IM8-9, SM1-3, IL6-8, ML1-8, SP2-5, IP1-3, MP1-3, HH1-10 | 2 | Aura → mouth and hand automatisms → mild combativeness & amnesia |
|  |  |  |  |  | 43 | IM1, IM8-9, SM1-3, IL6-8, ML1-8, SP2-5, IP1-3, MP1-3, HH1-10 | 2 | Aura |
| 2 | 45-50 | M | Bitemporal | Lamotrigine, Phenytoin, Valproic Acid | 90 | TP1-8, AM1-8, HB1-2, RA1-2 RH1-8, HH1-8 | 2 | Aura → postictal aphasia |
| | | | | | 120 | TP1-8, AM1-4, HB1-2 | 2 | Aura → postictal aphasia |
| 3 | 20-25 | F | Left Mesial Temporal | Trileptal, Vimpat | 120 | HH1-3, HB1-3, AM1-3, MH1-12, PH1-12, IA1-12, IM1-12, SA1-12, MA1-12 | 4 | Abdominal aura. |
|  |  |  |  |  | 120 | HH1-3, HB1-3, AM1-3, MH1-12, PH1-12, IA1-12, IM1-12, SA1-12, MA1-12 | 4 | Abdominal & gustatory aura |
| 4 | 30-35 | F | Right Mesial Temporal | Keppra, Lacosamide | 60 | HH2-3, EM8-9, HH1-12, HB1-12, TT1-12, OF1-12 | 4 | After stimulating AM3 with 50Hz, 4.6mA, 3s, patient felt "oozy" |
|  |  |  |  |  | 60 | AM1-2, EM9-10, HH1-12, HB1-12, TT1-12, OF1-12 | 4 | After stimulating AM4 with 5Hz, 7mA, 3 seconds, patient felt funny |

## 3. Results

To validate the use of the MaTiLDA interface for classifying complex brain states from persistent homology features from SEEG using various topological data analysis feature vectorizations and machine learning methods, we analyzed seizure events across eight seizures from four refractory epilepsy patients. We aimed to classify each of the seizure periods (characterized by aberrant activity spread to new regions) for each seizure using the NIC and MaTiLDA workflows as described above. For brevity, we present only the dimension 0 analyses here.

For four seizures, each seizure period had only one spread beyond seizure onset; thus, a series of binary classifications to compare onset and the first seizure period was conducted for each seizure. Due to space constraints, we review only the results for RF, SVM, and LR models using either the lifespan or persistence landscape vectorization methods. Model performance varied across all seizures, and no one algorithm or vectorization method outperformed others to consistently distinguish onset and seizure period one (***Table 2)***. We speculate that this is due to the imbalanced class sizes. For example, for patient YROB seizure two, only five seconds of the 120 second seizure were labeled as a part of seizure onset, making it very difficult for a learning algorithm to identify patterns in seizure onset. Receiver operating characteristic (ROC) curves can be seen for each of these models for all four seizures in ***Figure 4***.

**Table 2:** MaTiLDA’s output for model performance for RF, SVM, and LR models using lifespan or persistence landscape vectorization methods for four different seizures across two patients.

| Patient | Seizure | Algorithm | Vectorization | Accuracy | Precision | Recall | AUC |
| --- | --- | --- | --- | --- | --- | --- | --- |
| 1 | 1 | RF | Lifespan | 1 | 1 | 1 | 1 |
|  |  |  | Landscape | 0.9 | 1 | 0.67 | 0.83 |
|  |  | SVM | Lifespan | 0.9 | 1 | 0.67 | 0.83 |
|  |  |  | Landscape | 0.8 | 0.67 | 0.67 | 0.76 |
|  |  | LR | Lifespan | 0.8 | 1 | 0.33 | 0.67 |
|  |  |  | Landscape | 0.8 | 1 | 0.33 | 0.67 |
|  | 2 | RF | Lifespan | 0.67 | 0.5 | 0.33 | 0.58 |
|  |  |  | Landscape | 0.67 | 0.5 | 0.33 | 0.58 |
|  |  | SVM | Lifespan | 0.67 | 0.5 | 0.33 | 0.58 |
|  |  |  | Landscape | 0.67 | 0.5 | 0.33 | 0.58 |
|  |  | LR | Lifespan | 0.56 | 0 | 0 | 0.42 |
|  |  |  | Landscape | 0.56 | 0.5 | 0.33 | 0.58 |
| 2 | 1 | RF | Lifespan | 0.95 | 0.8 | 1 | 0.97 |
|  |  |  | Landscape | 1 | 1 | 1 | 1 |
|  |  | SVM | Lifespan | 0.8 | 0 | 0 | 0.5 |
|  |  |  | Landscape | 0.9 | 1 | 0.5 | 0.75 |
|  |  | LR | Lifespan | 0.8 | 0 | 0 | 0.5 |
|  |  |  | Landscape | 0.85 | 0.67 | 0.5 | 0.72 |
|  | 2 | RF | Lifespan | 0.96 | 0 | 0 | 0.5 |
|  |  |  | Landscape | 0.96 | 0 | 0 | 0.5 |
|  |  | SVM | Lifespan | 0.96 | 0 | 0 | 0.5 |
|  |  |  | Landscape | 0.96 | 0 | 0 | 0.5 |
|  |  | LR | Lifespan | 0.96 | 0 | 0 | 0.5 |
|  |  |  | Landscape | 0.96 | 0 | 0 | 0.5 |

The remaining four seizures extended three seizure periods beyond onset, resulting in a series of multiclass classifications. Due to space constraints, we limit our results to a discussion of RF models using the lifespans and persistence landscape vectorization methods. Again, no one algorithm or vectorization method outperformed others to consistently distinguish seizure periods, and there was high variation in model performance within and across seizures (***Figure 5***). Models typically showed better performance in situations with more balanced data.

**Figure 5:**
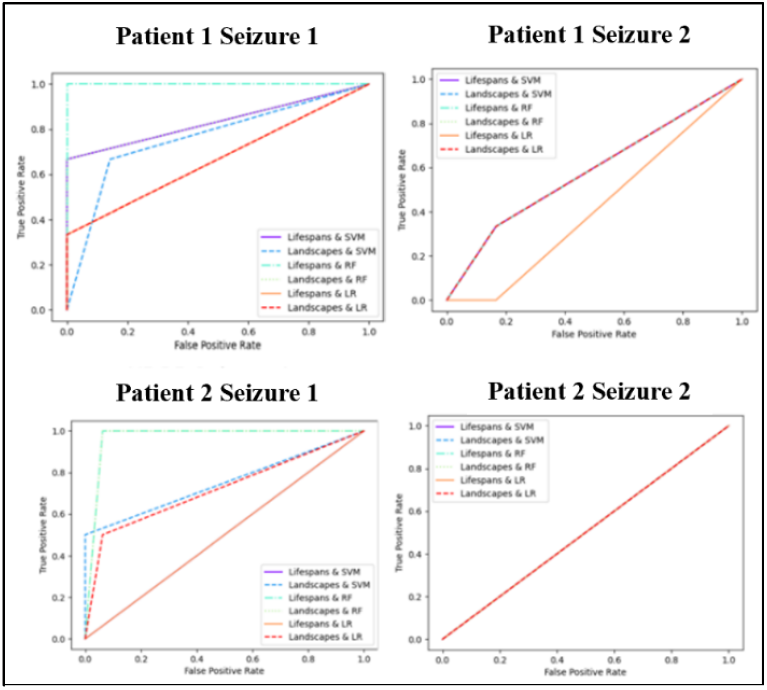
ROC curves for each seizure from the binary classification of seizure phase using lifespan or landscape vectors in SVM, RF, or LR.

**Figure 6:**
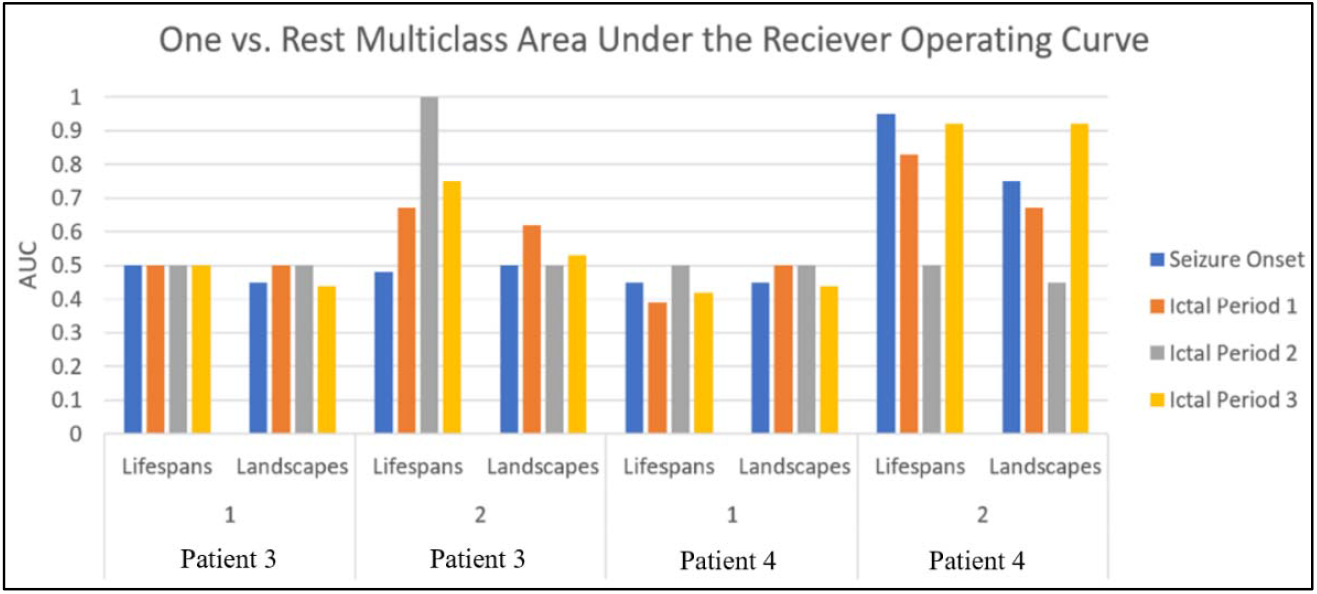
MaTiLDA’s One vs Rest AUC values for random forest seizure classification using lifespans or landscapes for each of the 4 multi-part seizures.

It is extremely difficult to quantify the changes in network topology during various ictal periods due to the complex interactions between brain regions during ictal periods. Our results show that for some seizures, automated machine learning analyses of topological data analysis may improve our ability to systematically quantify the brain network dynamics during ictal periods. Moreover, we have shown that MaTiLDA allows future research to further explore these methods for systematic quantification.

## 4. Discussion & Conclusion

The results of this evaluation demonstrate that MaTiLDA is an effective tool for analyzing complex topological features, enabling the detection of changes in brain networks during seizures. We have developed a novel pipeline that integrates topological data analysis (TDA) of stereotactic electroencephalography (SEEG) data with machine learning (ML) models. Our pipeline has is capable of accurately distinguishing brain states in several seizure phases using various common vectorizations of topological data and classical ML classification algorithms. Moreover, the platform provides a robust framework for quantitative comparison of different TDA vectorization methods and ML models in their ability to classify changes in dynamic brain networks. This study has contributed to the development of an accessible and reliable tool for TDA and ML analysis of SEEG data, with broad implications for neurological disorder diagnosis and treatment.

## Data Availability

All data produced in the present study are available upon reasonable request to the authors

